# Modified Robust Meta-Analytic-Predictive Priors for Incorporating Historical Controls in Clinical Trials

**DOI:** 10.1101/2023.01.28.23285146

**Authors:** Qiang Zhao, Haijun Ma

**Affiliations:** Gilead Sciences Inc, 333 Lakeside Dr, Foster City, CA 94404; Exelixis Inc, 1851 Harbor Bay Pkwy, Alameda, CA 94502

**Keywords:** historical control, dynamic borrowing, empirical Bayes, clinical trials, mixture distribution

## Abstract

Incorporating historical information in clinical trials has been of much interest recently because of its potential to reduce the size and cost of clinical trials. Data-conflict is one of the biggest challenges in incorporating historical information. In order to address the conflict between historical data and current data, several methods have been proposed including the robust meta-analytic-predictive (rMAP) prior method. In this paper, we propose to modify the rMAP prior method by using an empirical Bayes approach to estimate the weights for the two components of the rMAP prior. Via numerical calculations, we show that this modification to the rMAP method improves its performance regarding multiple key metrics.

## 1. Introduction

Randomized controlled trials (RCTs) have been used as the gold standard for years to establish the efficacy and safety of an investigational medical product. However, RCTs usually take a long time and are of high cost. Patient recruitment could be difficult for indications of rare diseases, pediatrics, and certain targeted sub-groups, which unduly delays development and market access of treatment for these indications. On the other hand, patient data from real world use such as registries, electronic health records, and historical clinical trials, have become more and more accessible. This has motivated the use of external historical data to enrich the control arm in RCTs.

Using historical data to aid RCTs is not a new idea (Pocock 1976). However, the regulatory agencies’ engagement in recent years has advanced this field considerably. FDA has released a series of guidance documents related to use of external and historical data in regulatory settings (FDA 2017; FDA 2019 a, b, c; FDA 2021). Use of historical controls is already quite common in medical device trials (CDRH 2021). EMA also published a draft guideline on registry-based studies to enhance the use of registry-based studies as a source of real-world evidence (EMA 2021).

One of the biggest statistical challenges in incorporating historical data is prior-data conflict which means that the true distribution of parameter of interest underlying the historical data could be different from that of the current control data. Dynamic borrowing methods have been proposed to handle the prior-data conflict, including the test-then-pool method (Viele et al. 2014), the power prior method (Ibrahim et al. 2000; Ibrahim et al. 2015), the commensurate prior method (Hobbs et al. 2011) and the meta-analytic-predictive (MAP) prior method (Neuenschwander et al. 2010).

In particular, this article focuses on the robust MAP prior method (Schmidli et al. 2014). The robust MAP (rMAP) prior is a mixture prior with two components – the first component derived from historical data and the second component to ensure robustness against prior-data conflict. However, it was not very clear how to assign weights to the two components in the mixture prior. In this paper, we propose to use an empirical Bayes (EB) method to derive the weight for the MAP prior, i.e. the component obtained from historical data, in the mixture rMAP prior. We organize this paper as follows. Section 2 delineates the proposed new method (EB-rMAP). Operating metrics based on the new method are evaluated for binomial data via numerical calculations and are compared with those based on competing methods in Section 3. The proposed method is applied to real clinical data in Section 4. We present the summary and discussions about the use of our method in Section 5.

## 2. Method

In clinical studies, binary outcomes, e.g. response to a study drug or not, are often of interest. In this paper, we consider a binary outcome *X*∼*Binom*(*n, θ*) with parameter *θ* ∈ [0, 1]. For example, *X* could be the number of responders observed out of a total of *n* subjects, and *θ* could be the true response rate to a study drug.

### 2.1 The Robust Meta-Analytic-Predictive (rMAP) Prior

(Neuenschwander et al. 2010) proposed using an MAP prior for incorporating historical data. As its name indicated, the MAP prior is estimated via meta-analysis of multiple historical studies. By applying the Bayes’ Theorem, the posterior distribution can then be derived and inferences will be conducted based on the posterior distribution.

(Schmidli et al. 2014) proposed to use a mixture of conjugate priors to approximate the MAP prior and to add a vague conjugate prior on top of the MAP prior to make it more robust, i.e.

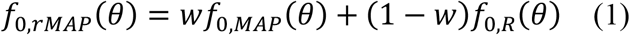

where *f*_0,*MAP*_(*θ*) is the MAP prior derived from historical data, *f*_0,*R*_(*θ*) is a vague prior, and *f*_0,*rMAP*_(*θ*) is the robust MAP prior. Moving forward in this article, we’ll use ‘MAP prior’ to denote the prior distribution based on historical data, i.e. *f*_0,*MAP*_(*θ*), while ‘rMAP prior’ refers to *f*_0,*rMAP*_(*θ*) in (1). The weight *w* impacts how quickly historical data are discounted with increasing prior-data conflict. In general, it was suggested that the weight *w* should be based on the degree of confidence in the relevance of the historical data (Schmidli et al. 2014). However, there was no clear guidance on how to quantify *w* yet.

It can be shown that in general the posterior distribution corresponding to the mixture prior distribution *f*_0,*rMAP*_(*θ*) is still a mixture of two distributions, however with different weights compared with those weights in the prior distribution:

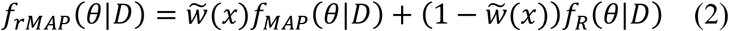

where *D* stands for observed data and *f*. (*θ*|*D*) is the posterior distribution corresponding to the prior distribution *f*_0_,. (*θ*). In general, 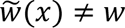. Using data that follows binomial distribution and its conjugate prior distribution as an example, it’s easy to show that

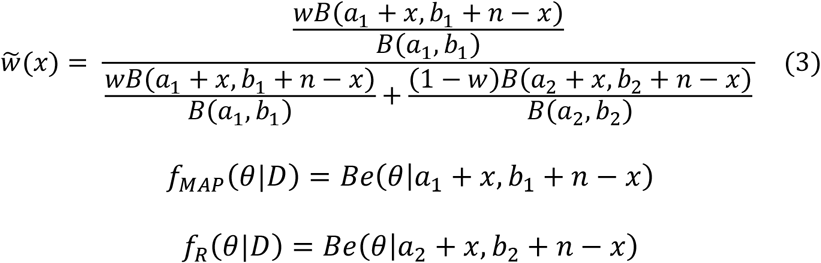

given a mixture prior distribution

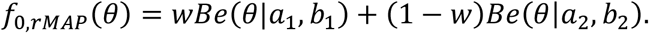

Here *Be*(*θ*|*a, b*) is the Beta distribution for *θ* with parameters *a* and *b*, and *B*(*a, b*) is the Beta function with parameters *a* and *b*. A common choice for *a*_2_ and *b*_2_ is that *a*_2_ = *b*_2_ = 1, i.e. the uniform distribution is selected as the vague prior *Be*(*θ*|*a*_2_, *b*_2_). Without loss of generalizability, we assume that *f*_0,*MAP*_(*θ*) in (1) consists of one Beta distribution, i.e. *Be*(*θ*|*a*_1_, *b*_1_). The above results can be easily extended to the case when the MAP prior consists of a mixture of Beta distributions (See Supplementary Material S1). It can be shown that when there’s large prior-data conflict, 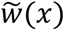 will be shrunk compared with its counterpart *w* in the mixture prior distribution (See Supplementary Material S1; Schmidli et al. 2014). In other words, the MAP prior will barely contribute to the posterior distribution in cases where there’s large prior-data conflict.

### 2.2 Empirical Bayes-based Robust MAP Prior (EB-rMAP)

In this paper, we propose to use an empirical Bayes method to quantify the weight *w* for the rMAP prior. In particular, assume that the estimate (e.g. the maximum likelihood estimate) of the parameter of interest based on the current control data is 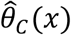, then the following probability is used as the weight of the MAP prior in the mixture prior in (1), i.e.

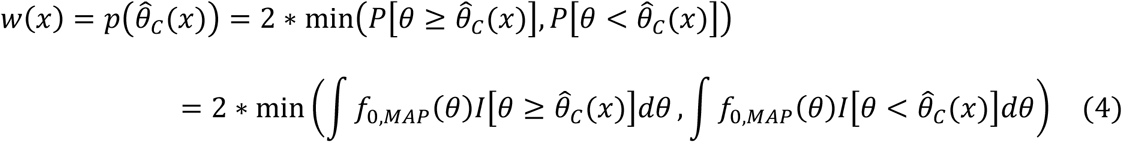

where *θ* denote the parameter of interest, and *f*_0,*MAP*_(*θ*) is the MAP prior distribution. By replacing *w* in (1) and (2) with 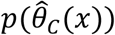, we get a modified robust MAP prior *f*_0,*EB*−*rMAP*_(*θ*) and the corresponding posterior distribution *f*_*EB*−*rMAP*_(*θ*|*D*),

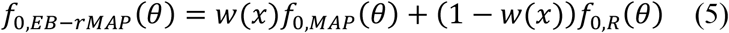

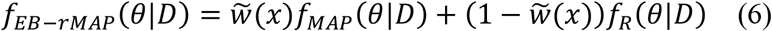

Given a mixture of Beta prior distributions, *f*_0,*EB*−*rMAP*_(*θ*) = *w*(*x*)*Be*(*θ*|*a*_1_, *b*_1_) + (1 − *w*(*x*))*Be*(*θ*|*a*_2_, *b*_2_),

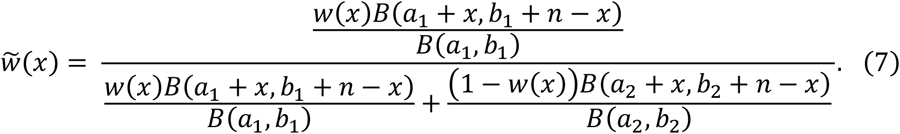

Notice that the only difference between (7) and (3) is that *w*(*x*) in (7) is a function of observed data as specified by (4) whereas *w* was a fixed number in (3).

The idea behind this EB approach is similar to p-value used in the hypothesis testing, where p-value measures how likely observing more extreme data than the currently observed data is, given the null hypothesis is true. In other words, p-value indicates how similar the distribution behind the current data to the null hypothesis distribution. Notice that *w*(*x*) is calculated in the same way as p-value is calculated in a 2-sided test if the MAP prior was taken as the distribution under the null hypothesis. So, similar to p-value, *w*(*x*) or 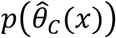 measures the similarity between the current observed data and the MAP prior distribution. A small *w*(*x*) or 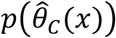 indicates that the parameter distribution behind the current observed data is dissimilar to the MAP prior distribution. This is the rationale that 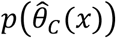 is used as the weight for the MAP prior in the mixture prior distribution.

## 3. Operating Metrics

We evaluated the performance of the EB-rMAP method and did comparisons with the rMAP method with different fixed *w* using the example historical data in (Viele et al. 2014) where there’re 65 responders among 100 patients. Accordingly, the beta distribution with parameters 65 and 35, i.e. Be(65, 35), was selected as *f*_0,*MAP*_. The Uniform distribution, or Be(1, 1) was used as *f*_0,*R*_ in (1) and (4). In other words, *a*_1_ = 65, *b*_1_ = 35, *a*_2_ = 1, *b*_2_ = 1 in the mixture prior distribution through Section 3. In the current study, assume that there’re two arms, one control arm and one treatment arm, each with sample size *n* =100. The number of responders obtained in current control arm and current treatment arm are models as,

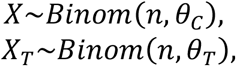

where *θ*_*C*_ and *θ*_*T*_ are true response rate in current control arm and current treatment arm, respectively. The objective of this study is to compare *θ*_*T*_ with *θ*_*C*_ to determine whether the treatment has higher response rate than the control, with the null hypothesis *H*_0_: *θ*_*T*_ ≤ *θ*_*C*_. Let *θ*_*C*_ varies from 0.5 to 0.85.

Prior distributions for each method are list out in table 1.

**Table 1.** Weight in prior distributions

| | Prior Distribution $f_0(\theta_C)$ | $w$ |
| --- | --- | --- |
| MAP | $wBe(\theta_C\vert a_1, b_1) + (1 - w)Be(\theta_C\vert a_2, b_2)$ | 1 |
| rMAP | | given fixed $w \in [0, 1]$ |
| EB-rMAP | $w(x)Be(\theta_C\vert a_1, b_1) + (1 - w(x))Be(\theta_C\vert a_2, b_2)$ | $w(x)$ defined by (4) |

### 3.1 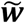 in the posterior distribution

By (3) and (7), 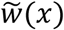 for different methods are shown in table 2.

**Table 2.**
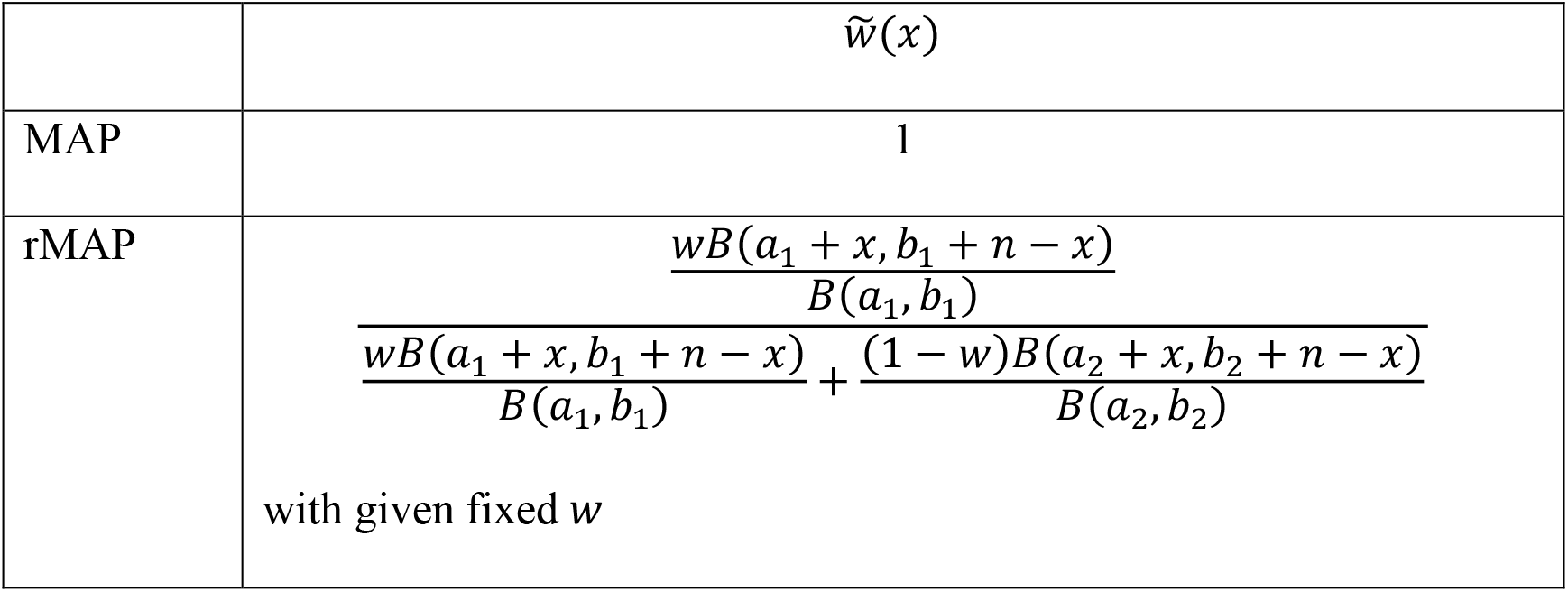
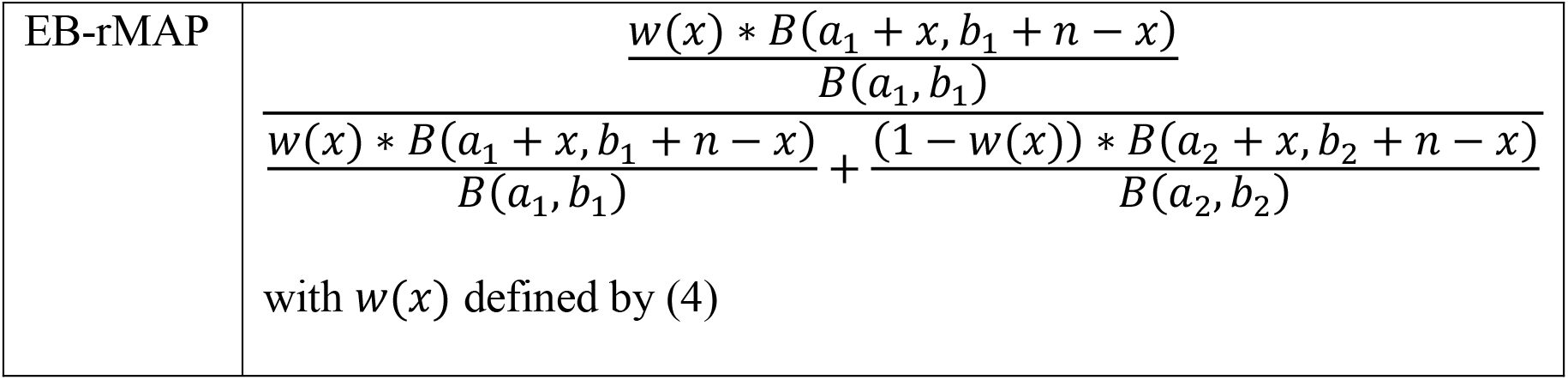
Weight in posterior distributions

The mean of 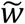 is calculated as

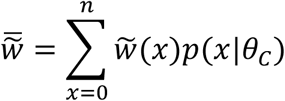

where *p*(*x*|*θ*_*C*_) refers to the binomial probability mass function. The relationship between 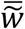 and true *θ*_*C*_ for different methods are shown in Figure 1. For the MAP method without robustification, 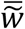 always equal to 1. For the rMAP and the EB-rMAP method, 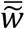 decreases when the true parameter *θ*_*C*_ differs from 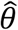, which is 0.65 in this case, based on historical data. In particular, when *θ*_*C*_ = 0.8 on average 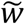 can still be as high as 0.61 given *w*=0.8. Considering that the true parameter *θ*_*C*_=0.8 indicates substantial data conflict between current data and historical data 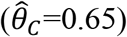, assigning a weight as high as 0.61 to the historical data seems too high. On the contrary, EB-rMAP has 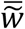 as low as 0.03 on average. When *θ*_*C*_ = 0.65, i.e. no data conflict, the 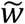 is around 0.73 on average based on EB-rMAP. This is not as high as the 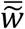 based on rMAP with *w*=0.8, which indicates that EB-rMAP is relatively conservative in borrowing historical information. However, the conservativeness of the EB-rMAP method provides safeguard against borrowing too much from historical data when there’s data conflict as discussed above.

**Figure 1.**
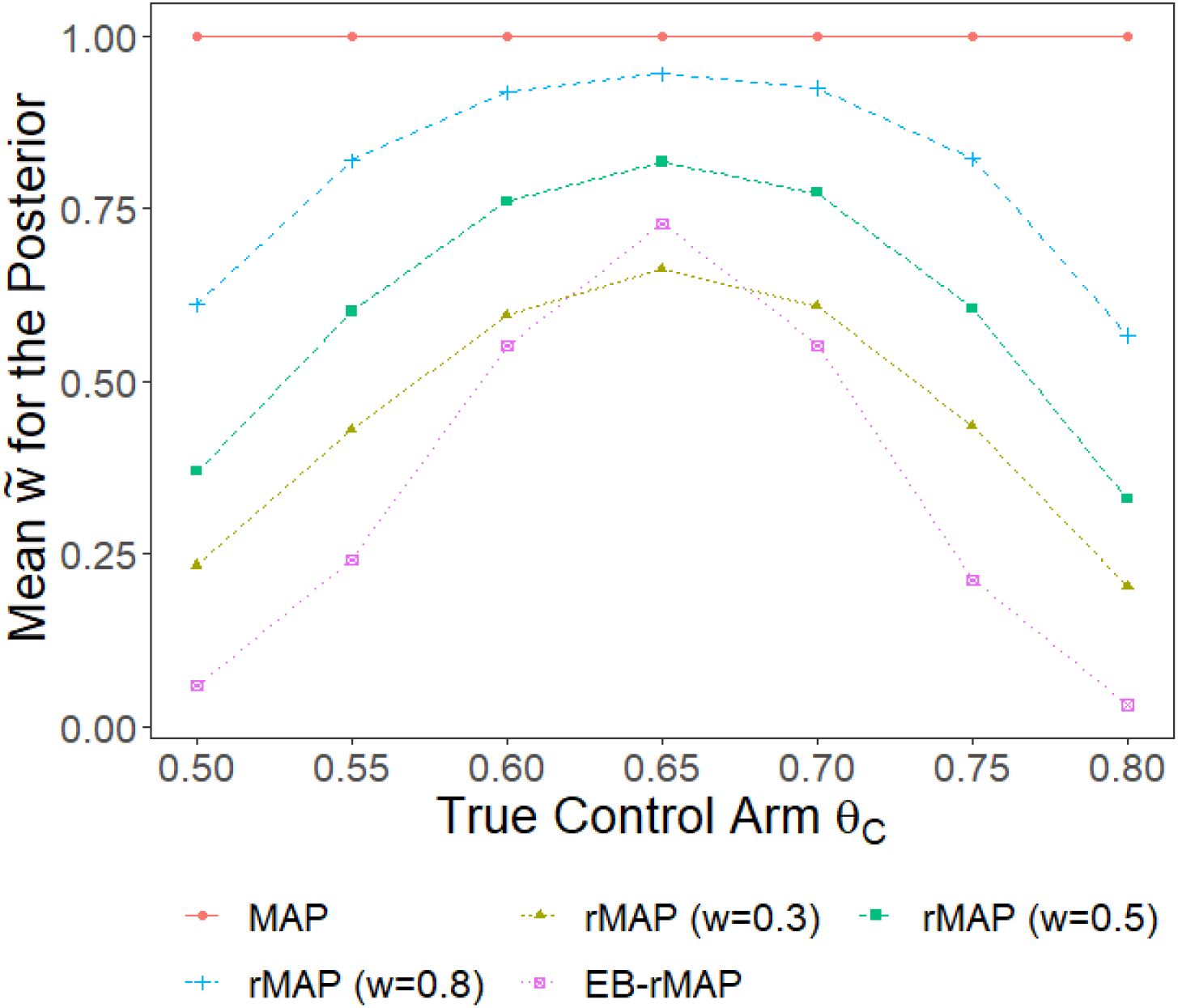
Weight in the posterior distribution based on MAP, rMAP, and EB-rMAP methods for historical data 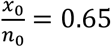

### 3.2 Effective historical sample size

Effective historical sample size (EHSS) is an important metric to measure how much information is borrowed from historical data. In this article, EHSS is calculated by comparing the precision of the parameter estimates before and after incorporating historical data (Hobbs et al. 2013), i.e.

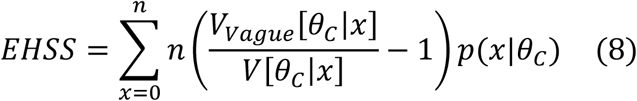

where *n* is the sample size of the control arm in the current study, *V*_*Vague*_[*θ*_*C*_|*x*] is the variance of *θ*_*C*_ regarding a posterior distribution derived based on a vague prior e.g. Be(0.01, 0.01), and *V*[*θ*_*C*_|*x*] refers to the variance of *θ*_*C*_ regarding to the posterior distributions derived based on priors listed in table 1 for different methods. More specifically, as the observed number of responders *X* in the control arm follows a binomial distribution, i.e. *X*∼*Binom*(*n, θ*_*C*_), the posterior distribution for *θ*_*C*_ is *Be*(*x* + 0.01, *n* − *x* + 0.01). It follows that

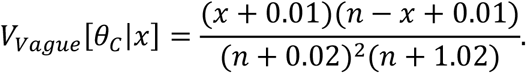

Since usually *n* ≫ 0.01,

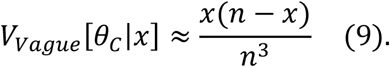

As for *V*[*θ*_*C*_|*x*], the posterior distribution for *θ*_*C*_ is a mixture of two other distributions as shown in (2). It’s easy to show (See Supplementary Material S2) that

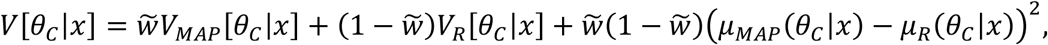

where *μ*_*MAP*_(*θ*_*C*_|*x*), *μ*_*R*_(*θ*_*C*_|*x*), *V*_*MAP*_[*θ*_*C*_|*x*] and *V*_*R*_[*θ*_*C*_|*x*] are the mean and variance of *θ*_*C*_ regarding the posterior distribution *f*_*MAP*_(*θ*_*C*_|*x*) and *f*_*R*_(*θ*_*C*_|*x*), respectively.

Figure 2(A) shows the EHSS for each method. Note that as EHSS’s are estimated, they could be negative. Using the MAP method, EHSS is around 100 when *θ*_*C*_ is less than 0.65 and decreases to 58 when *θ*_*C*_ increases to 0.85. This decrease trend is driven by the numerator *V*_*Vague*_[*θ*_*C*_|*x*] of (8), which achieves its maximum when *θ*_*C*_=0.5 (see (9)) and decreases when *θ*_*C*_ shifts away from 0.5. Unlike the MAP method which resulted in a maximum EHSS when *θ*_*C*_=0.55, both the rMAP method and the EB-rMAP method achieve its maximum EHSS when *θ*_*C*_=0.65, i.e. when current data is congruent with the historical data. When *θ*_*C*_ differs from the observed 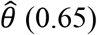 (0.65) based on historical data, EHSS’s based on both the rMAP method and the EB-rMAP method decrease. This illustrates the robustness of these two methods in handling data conflict. More specifically, when *θ*_*C*_ = 0.65 EHSS is approximately 70 using the EB-rMAP method. This is close to the EHSS of using the rMAP method with *w* = 0.5.

**Figure 2.**
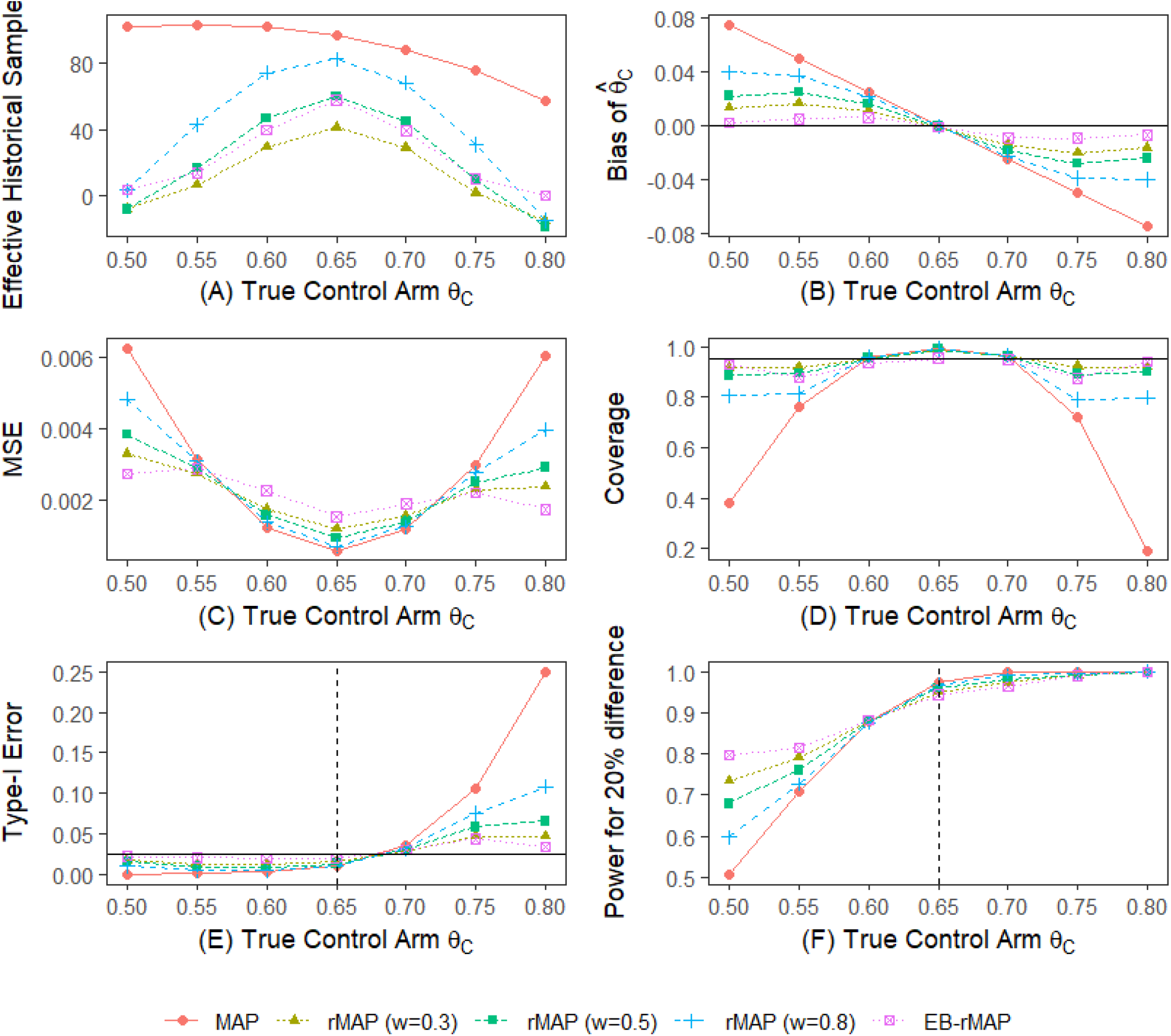
Performance metrics of MAP, rMAP, and EB-rMAP methods for historical data 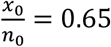 and varying true parameter *θ*_*C*_. (A) Effective historical sample size. (B) Bias. (C) Mean squared error. (D) Coverage. (E) Type I error. (F) Power to detect 0.2 difference in response rate.

### 3.3 Bias

By definition,

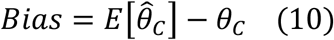

In Bayesian framework, 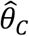 is taken as the mean of the posterior distribution of *θ*_*C*_, i.e.

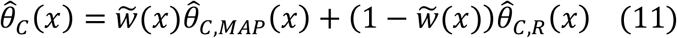

where 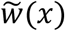 takes values shown in table 2 for each method. In cases where observed data follows Binom(*n, θ*) distribution, and *f*_0_(*θ*) = *w* * *Be*(*θ*|*a*_1_, *b*_1_) + (1 − *w*) * *Be*(*θ*|*a*_2_, *b*_2_), then

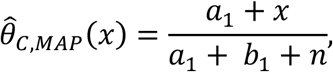

and

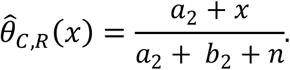

Given the probability mass function *p*(*x*|*θ*_*C*_) for observed data,

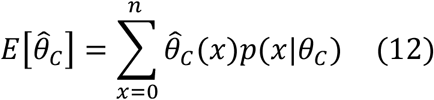

By substituting (11) and 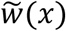 in table 2 into (12), 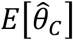 can be calculated for each method. Bias for each method is then calculated by applying (10) and is plotted in figure 2(B). It can be seen that compared with other methods, the EB-rMAP method results in smaller bias consistently across wide range of true *θ*_*C*_.

### 3.4 Mean squared error (MSE)

By definition,

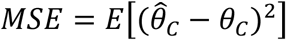

Similar to 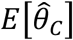, given the probability mass function *p*(*x*|*θ*_*C*_) for observed data,

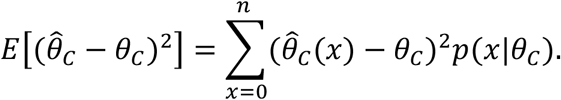

Figure 2(C) shows the MSE versus the true *θ*_*C*_ for different methods. For the EB-rMAP method, MSE is relatively larger than other methods mainly due to the variability introduced by making the weight *w* in the prior distribution a random variable instead of a fixed number. However, EB-rMAP controls the MSE level better when there’s data conflict, e.g. when *θ*_*C*_ is below 0.55 or above 0.75 compared with other methods because of the smaller bias of using the EB-rMAP method.

### 3.5 Type-I error and power

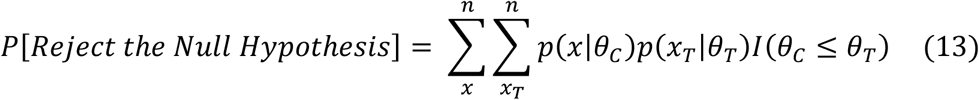

where *I*(*θ*_*C*_ ≤ *θ*_*T*_) = 1 when *θ*_*C*_ ≤ *θ*_*T*_ or 0 otherwise. For ease of calculations, *I*(*θ*_*C*_ ≤ *θ*_*T*_) is approximated by randomly generating 1000 data points from the posterior distribution of *θ*_*C*_ and *θ*_*T*_ given observed *x* and *x*_*T*_. The posterior distribution for *θ*_*T*_ was obtained by assuming a vague prior *Be*(0.01, 0.01).

Type-I error and power are then calculated by substituting *θ*_*T*_ with *θ*_*C*_ and *θ*_*C*_ + 0.2 respectively into (13). Figure 2(E) and 2(F) illustrate the type-I errors and powers using different methods. Due to the bias introduced by historical data, type-I errors based on the MAP method, the rMAP method and the EB-rMAP method are all below the nominal level 0.025 when the true *θ*_*C*_ is below 0.65. Type-I errors could be above 0.025 when the true *θ*_*C*_ is above 0.65. However, there’s clear decrease in type-I error using the EB-rMAP method when the true *θ*_*C*_ is beyond 0.75, which shows the control of type-I error when using this method. On the contrary, the MAP method and the rMAP method show less control over type-I error. Especially when the true *θ*_*C*_ is 0.8, type-I error based on the MAP method could be as high as 0.25, the type-I error based on the rMAP method with *w*=0.8 could be as high as 0.1.

Similar to the trend of type-I errors, powers using all three methods are below 90% when the true *θ*_*C*_ is below 0.6, with the EB-rMAP method gives the highest power across all methods considered. When the true *θ*_*C*_ is above 0.6, the powers using all three methods are above 90%, with the EB-rMAP method gives relatively lower power. These observations are consistent with the relative smaller bias of using the EB-rMAP method across the whole range of the true *θ*_*C*_ considered.

### 3.6 Coverage probability

Coverage probability of a 95% credible interval for *θ*_*C*_ is calculated as

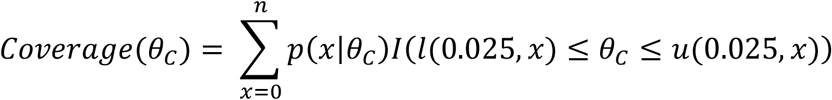

where *l*(0.025, *x*) and *u*(0.025, *x*) are the lower and upper 2.5% percentile of the posterior distribution of *θ*_*C*_. Figure 2(D) compares the coverage probabilities of the 95% credible intervals based on different methods. The common trend observed is that coverage probability is higher when *θ*_*C*_ is around the observed 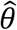, i.e. 0.65 based on historical data. This is because commensurate historical data enhances the precision of the estimate of *θ*_*C*_. Coverage probabilities then decrease when *θ*_*C*_ shifts away from 0.65 due to the increased bias caused by data conflict. However, the coverage probability based on EB-rMAP will go back to around 0.95 when *θ*_*C*_ approaches 0.5 or 0.85, whereas coverage probabilities based on MAP or rMAP with *w* = 0.8 could be as low as below 0.4 or 0.8, respectively. This again shows that the EB-rMAP method is more robust than other the MAP method and the rMAP method.

## 4. Application to Clinical Studies

To illustrate the application of the EB-rMAP method, we used 2 clinical trials (Rubinstein et al. 2001; Wunderink et al. 2003; and Gravestock et al. 2017) comparing two drugs for the treatment of nosocomial pneumonia as an example. Table 3 shows the results of the 2 trials for cure rate in the clinically evaluable subset of patients.

**Table 3.** Cure Rates in Example Clinical Studies

| Study | Cure rate |
| --- | --- |
| Rubinstein (2001) | 62/91 (68.1%) |
| Wunderink (2003) | 111/171 (64.9%) |

The earlier study (Rubinstein 2001) is selected as the historical study, and the (Wunderink 2003) study is selected as the ‘current’ study. As it’s observed that 62 patients were cured out of the total 91 patients in the historical study, the Beta(62, 29) distribution is chosen as the prior distribution (*f*_0,*MAP*_(*θ*)) based on historical data.

In the current study, there’re 111 patients cured out of the total 171 patients. So, the estimated cure rate is 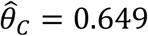. By plugging this into (4), we get

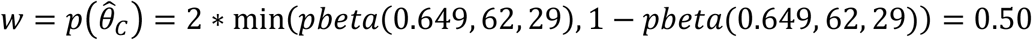

where *pbeta*(*x, a, b*) is the cumulative probability function for the Beta(a, b) distribution. Let *f*_0,*R*_(*θ*) = *Beta*(1, 1) as suggested in (Schmidli et al. 2014), then by (5)

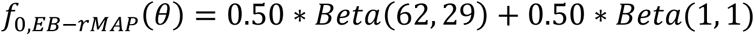

By (6) and (7), the posterior distribution is

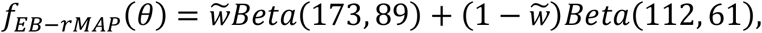

where

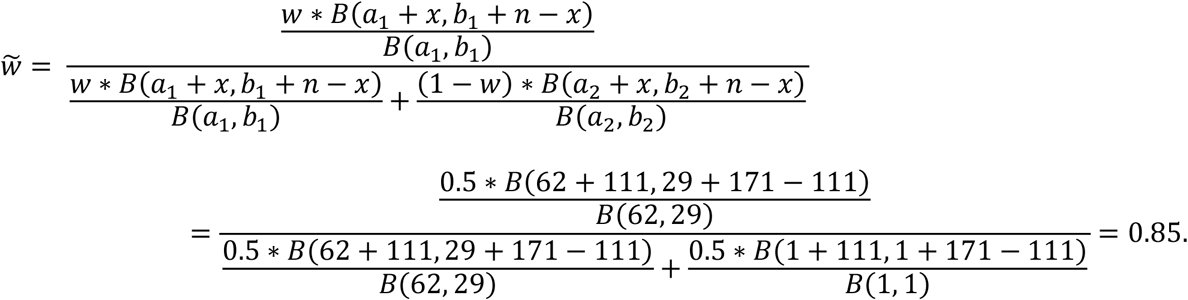

Inferences about parameter *θ* can then be performed based on its posterior distribution *f*_*EB*−*rMAP*_(*θ*) = 0.85 * *Beta*(173, 89) + 0.15 * *Beta*(112, 61).

Since the effective historical sample size, *EHSS*(*x*) depends on the current observed data, a natural application of the EB-rMAP method is in adaptive designs, where the interim data from a current study can be used to estimate *EHSS*(*x*), and the sample size for the post-interim part of the current study can be adjusted according to *EHSS*(*x*). More specifically, at the interim analysis, by applying the observed data into (8), the effective sample size (*EHSS*(*x*)) contributed by the historical data can be calculated. Assume *n*_*C*_ was the original planned sample size for the control arm, then the total sample size for the control arm can be reduced to *n*_*C*_ − *EHSS*(*x*). Depending on the sample size *n*_*C,IA*_ in the control arm by interim analysis, the post-interim sample size for the control arm can be calculated as max(0, *n*_*C*_ − *EHSS*(*x*) − *n*_*C,IA*_).

## 5. Discussions

Robust meta-analytic-predictive priors provide a convenient approach to borrow information from historical control data while taking care of data-conflict between historical control data and current control data. However, it was not clear how much weight should be assigned to each component in the robust MAP priors. In this article, we borrowed the idea of p-value in the hypothesis testing setup and proposed an EB approach to estimate this weight by comparing the parameter estimate based on current data versus the MAP prior distribution for the parameter of interest.

On the one hand, this EB-rMAP method complemented the original rMAP method by providing an interpretable choice for the weight parameter in the mixture prior distribution. More importantly, we have shown that using the EB-rMAP method, the bias caused by data conflict can be further reduced compared with using the robust MAP priors or the MAP priors. As a result, type-I errors are better controlled using the EB-rMAP method. A downside of using this EB approach is that when the data-conflict is subtle, the EB-rMAP method resulted in relatively larger MSE. This is caused by extra variability introduced by making the weight vary along with observed data instead of fixed. Considering that controlling type-I errors is of most concern in borrowing historical data in a regulatory setting and the power is comparable or higher using the EB-rMAP method compared with using other competing methods, this tradeoff in larger MSE is acceptable.

It’s worth to notice that the proposed p-value like approach is not the unique choice of the weight for the rMAP prior. However, weight in our proposed method shares a similar interpretation as that of the well-known p-value concept, so it would be easier to be accepted by a wider group of practitioners. Compared with other methods, e.g. the elastic MAP prior method (Jiang et al. 2021), our method is more straightforward and computationally less complex.

In addition, in this article the EB-rMAP method was illustrated using binary endpoint that follows binomial distribution. However, it is readily generalizable to endpoints follow other distributions e.g. normal distribution.

## Data Availability

All data produced in the present study are available upon reasonable request to the authors

## Supplementary Materials

### S1

Assume *X*∼*Binom*(*n*; *θ*); prior distribution for *θ* is *f*_0_(*θ*) = *wBe*(*θ*|*a*_1_, *b*_1_) + (1 − *w*)*Be*(*θ*|*a*_2_, *b*_2_), then posterior distribution for *θ* will be:

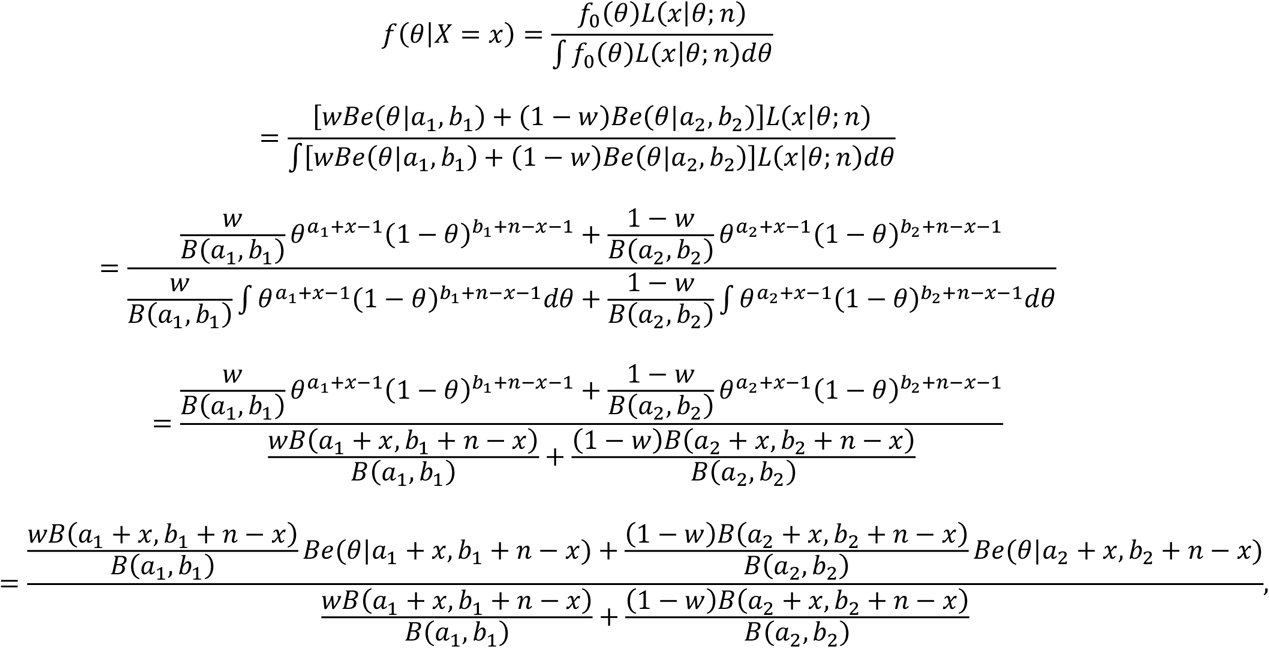

where

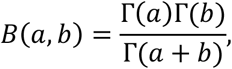

and *Be*(*θ*|*a, b*) is the Beta distribution with parameters *a* and *b* for *θ*.

Let

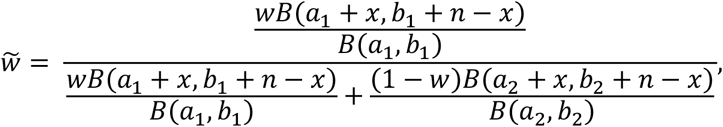

then the posterior distribution of *θ* can be re-written as

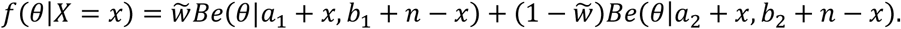

As an example, given *n* = 30, *a*_1_ = 65, *b*_1_ = 35, *a*_2_ = 1, *b*_2_ = 1, *w* = 0.3, 0.5, and 0.8, plot of 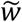 versus *x*/*n* is as below.

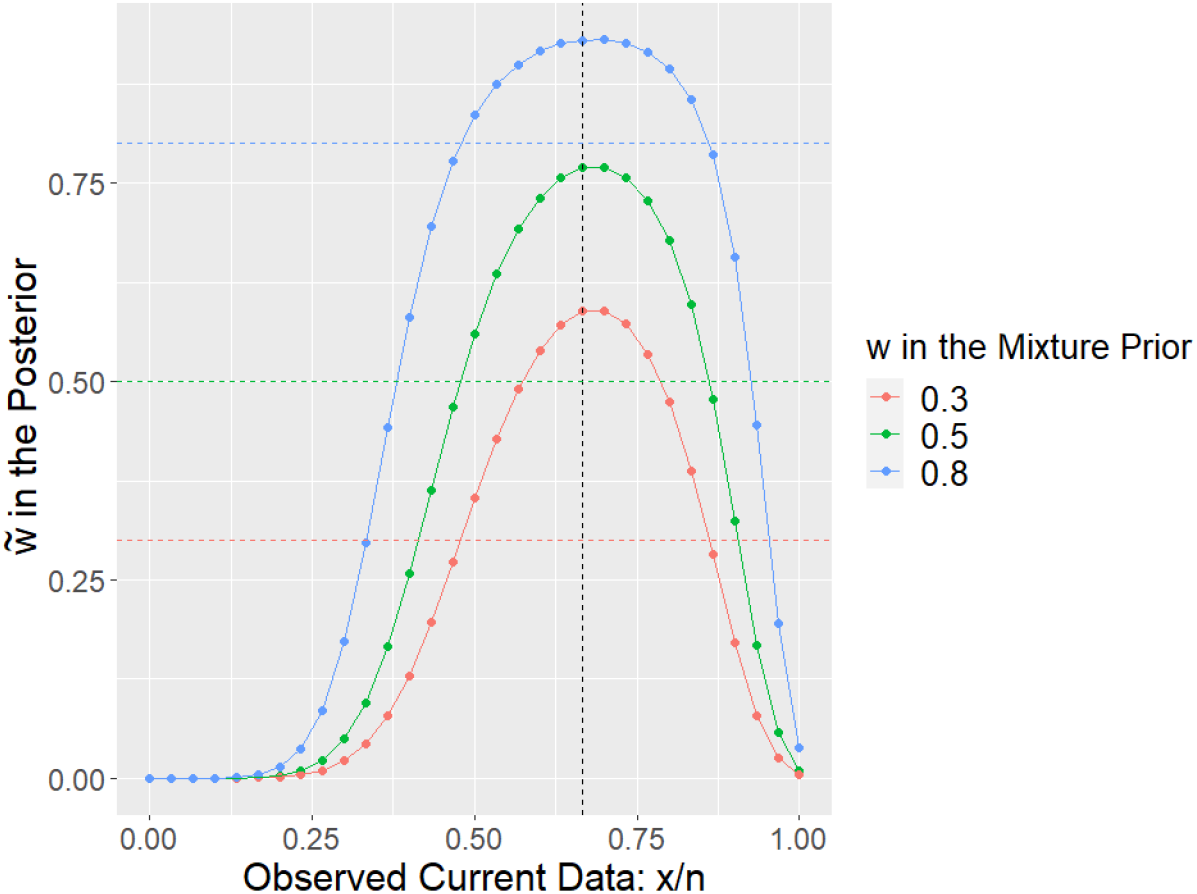

In general, when the prior distribution is a mixture of K (K ≥ 2) distributions, e.g. 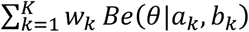, then

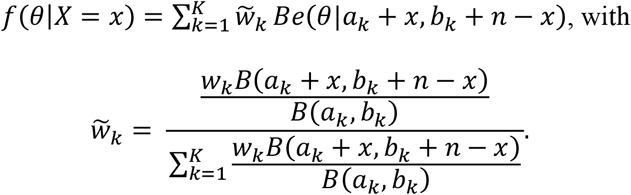

### S2

By (2) in main body of this article,

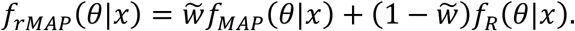

By linearity of expectations,

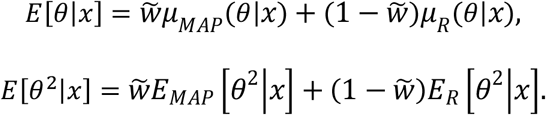

By definition *V*[*θ*|*x*],

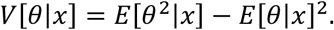

Substitute *E*[*θ*|*x*] and *E*[*θ*^2^|*x*] into the above formula,

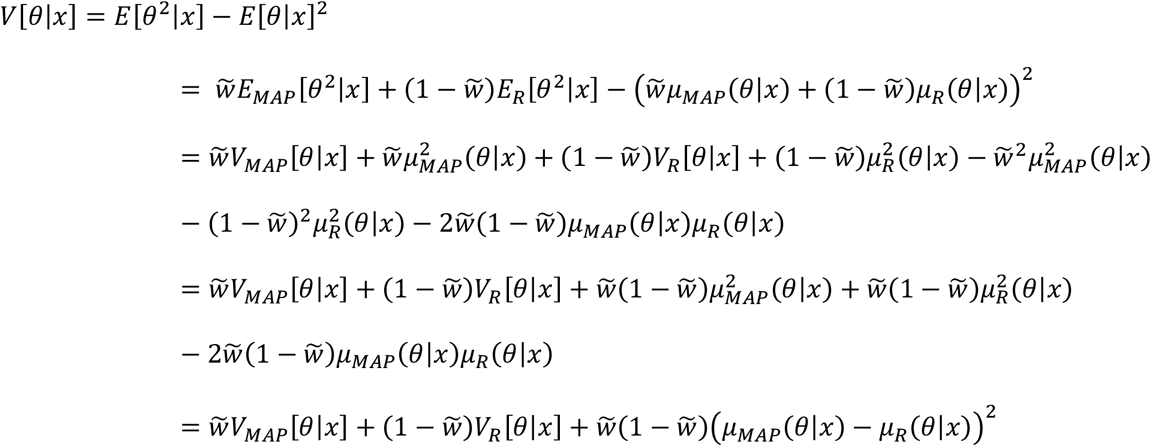

When *θ* = *θ*_*C*_,

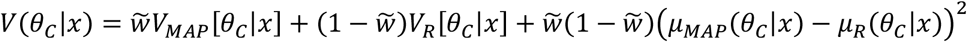

In general, when

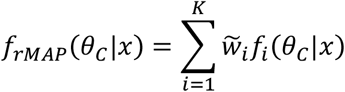

with 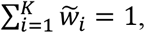,then,

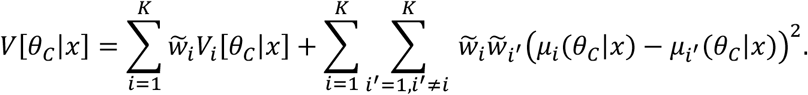

## References

Center for Devices and Radiological Health (CDRH), Examples of Real-World Evidence (RWE) Used in Medical Device Regulatory Decisions. https://www.fda.gov/media/146258/download 2021.

European Medicines Agency (EMA), Guideline on registry-based studies, 2021.

FDA Guidance for Industry and Food and Drug Administration Staff: Use of Real-World Evidence to Support Regulatory Decision-Making for Medical Devices, 2017.

FDA Guidance for Industry: Rare diseases: common issues in drug development. https://www.fda.gov/media/120091/download. February 2019 (Revision 1).

FDA Guidance for Industry: Rare diseases: natural history studies for drug development. https://www.fda.gov/media/122425/download. March 2019.

FDA Guidance for Industry: Demonstrating substantial evidence of effectiveness for human drug and biological products. https://www.fda.gov/media/133660/download. December 2019.

FDA Guidance for Industry: Considerations for the Use of Real-World Data and Real-World Evidence to Support Regulatory Decision-Making for Drug and Biological Products, 2021.

Gravestock, I., Held, L., & COMBACTE-Net consortium. (2017). Adaptive power priors with empirical Bayes for clinical trials. Pharmaceutical Statistics, 16(5), 349–360.

Hobbs, B. P., Carlin, B. P., Mandrekar, S. J., & Sargent, D. J. (2011). Hierarchical commensurate and power prior models for adaptive incorporation of historical information in clinical trials. Biometrics, 67(3), 1047–1056.

Hobbs, B. P., Carlin, B. P., & Sargent, D. J. (2013). Adaptive adjustment of the randomization ratio using historical control data. Clinical Trials, 10(3), 430–440.

Ibrahim, J. G., & Chen, M. H. (2000). Power prior distributions for regression models. Statistical Science>, 46–60.

Ibrahim, J. G., Chen, M. H., Gwon, Y., & Chen, F. (2015). The power prior: theory and applications. Statistics in medicine, 34(28), 3724–3749.

Jiang, L., Nie, L., & Yuan, Y. (2021). Elastic priors to dynamically borrow information from historical data in clinical trials. Biometrics>.

Neuenschwander, B., Capkun-Niggli, G., Branson, M., & Spiegelhalter, D. J. (2010). Summarizing historical information on controls in clinical trials. Clinical Trials>, 7, 5–18.

Pocock, S. J. (1976). The combination of randomized and historical controls in clinical trials. Journal of chronic diseases, 29(3), 175–188.

Rubinstein, E., Cammarata, S. K., Oliphant, T. H., & Wunderink, R. G. (2001). Linezolid versus vancomycin in the treatment of patients with nosocomial pneumonia: a randomized, double-blind, multicenter study. Clin Infect Dis, 32(3), 402–142.

Schmidli, H., Gsteiger, S., Roychoudhury, S., O’Hagan, A., Spiegelhalter, D., & Neuenschwander, B. (2014). Robust meta-analytic-predictive priors in clinical trials with historical control information. Biometrics>, 70(4), 1023–1032.

Viele, K., Berry, S., Neuenschwander, B., Amzal, B., Chen, F., Enas, N., … & Thompson, L. (2014). Use of historical control data for assessing treatment effects in clinical trials. Pharmaceutical statistics, 13(1), 41–54.

Wunderink, R. G., Cammarata, S. K., Oliphant, T. H., Kollef, M. H., & Linezolid Nosocomial Pneumonia Study Group. (2003). Continuation of a randomized, double-blind, multicenter study of linezolid versus vancomycin in the treatment of patients with nosocomial pneumonia. Clinical therapeutics, 25(3), 980–992.

